# Preparing for future Shigella vaccine introduction in Pakistan: a multi-stakeholder qualitative study of policy readiness and implementation

**DOI:** 10.64898/2026.07.31.26359393

**Authors:** Naveed Ahmed, Nasreen Mithani, Zubair Latif, Irum Fatima, Irfan Ghaffar, Mohammad Tahir Yousafzai, Farah Naz Qamar

**Author notes:** **Corresponding author:** Farah Naz Qamar, Department of Paediatrics and Child Health Aga Khan University, **Co-corresponding author:** Mohammad Tahir Yousafzai, Department of Paediatrics and Child Health Aga Khan University.

## Abstract

**Background:** Shigella remains an important cause of childhood diarrhoeal disease and a growing contributor to antimicrobial resistance in low- and middle-income countries (LMICs). Several Shigella vaccines are in development, yet little is known about community and stakeholder perspectives regarding the introduction of future Shigella vaccines in Pakistan. We explored perceptions related to diarrhoeal disease, vaccine acceptance, and health system readiness to inform the future introduction of a Shigella vaccine.

**Methods:** We conducted a qualitative study in Karachi, Pakistan, between August 2024 and March 2025. Data collection comprised six focus group discussions (FGDs) with caregivers of children under five years of age, and 16 in-depth interviews (IDis) with key stakeholders including healthcare providers, Expanded Programme on Immunization (EPI) officials, policymakers, technical experts, school administrators, and community and religious leaders. Semi-structured interview guides informed by the Health Belief Model. Audio-recorded interviews were transcribed verbatim, translated into English, and analyzed thematically using both deductive approaches and inductive approaches.

**Results:** Four interconnected themes emerged from the data: perceptions of diarrhoeal disease and Shigella; factors shaping acceptance of a future Shigella vaccine; evidence and health-system readiness for vaccine introduction; and vaccine attributes, adoption, and sustainability. Although childhood diarrhoeal disease was widely recognized, awareness of Shigella as a specific pathogen was limited. Participants expressed conditional acceptance of a future Shigella vaccine, influenced by trust, perceived safety, and recommendations from healthcare providers and community leaders.

Stakeholders emphasized the need for stronger local evidence on disease burden and cost effectiveness to support vaccine prioritization, while identifying Pakistan’s existing immunization infrastructure as a strength for implementation. Some participants favoured combination vaccines to reduce injections and simplify vaccine delivery, while others emphasized that vaccination should complement continued improvements in water, sanitation, and hygiene.

**Conclusion:** Knowledge of Shigella and its vaccine was limited, but stakeholders supported future vaccine introduction if informed by local evidence, policy commitment, and program integration.

**What is already known on this topic:** Shigella remains a leading cause of childhood diarrhoeal disease and antimicrobial resistance in low and middle-income countries, and several vaccine candidates are in advanced stages of development. However, little is known about stakeholder perspectives, policy readiness, and health-system preparedness for the introduction of a future Shigella vaccine in Pakistan.

**What this study adds:** This study provides the first multi-stakeholder qualitative assessment of perceptions of a future Shigella vaccine in Pakistan. While awareness of Shigella was limited, stakeholders considered vaccine introduction feasible within the existing immunization programme, provided that stronger locally generated evidence, sustained policy commitment, and early implementation planning support decision-making.

**How this study might affect research, practice or policy:** The findings highlight the need to complement epidemiological evidence with early stakeholder engagement and implementation planning before vaccine introduction. Strengthening surveillance, generating nationally relevant evidence, and leveraging Pakistan’s established Expanded Programme on Immunization (EPI) and trusted community networks may facilitate successful introduction of a future Shigella vaccine.

## Introduction

Diarrhoeal diseases remain a leading cause of morbidity and mortality among children under five years of age globally, accounting for an estimated 1.7 billion episodes and more than 400,000 deaths each year (1). The burden is disproportionately concentrated in low- and middle-income countries, where limited access to safe water, sanitation, and timely health care continues to drive high transmission and recurrent illness (2). Despite improvements in child survival and the expansion of routine immunization programmes, diarrhoeal disease continues to place a substantial burden on health systems and households through increased healthcare utilization, caregiver productivity loss, and out-of-pocket expenditure, particularly in resource-limited settings (3, 4).

Among bacterial causes of diarrhoea, Shigella is a leading cause of moderate-to-severe illness in young children and accounted for an estimated 63,713 deaths among children under five years of age in 2016 (5–8). Beyond acute illness, Shigella infection has been associated with recurrent diarrhoea, growth faltering, and adverse developmental outcomes (9, 10). Its public health importance is further amplified by the rapid emergence of antimicrobial resistance, which has reduced the effectiveness of commonly used antibiotics and limited treatment options in many endemic regions, including South Asia (5, 11).

Pakistan remains a high-burden setting for childhood diarrhoeal disease, characterized by persistent challenges in water, sanitation, and hygiene, high healthcare utilization for paediatric diarrhoea, and considerable socioeconomic vulnerability among affected households (12). Surveillance initiatives, including the Global Enteric Multicenter Study (GEMS) and the Enterics for Global Health (EFGH) Shigella Surveillance Study, have generated important epidemiological, microbiological, and economic evidence on Shigella burden in Pakistan and other endemic settings (5, 13, 14). These data have strengthened the evidence base for Shigella vaccine development and future policy decisions (13, 15).

However, for the epidemiological evidence to translate into vaccine policy, a persistent know-do gap, whereby scientific evidence is insufficiently integrated into decision-making, planning, and implementation processes (16, 17). In Pakistan’s decentralized health system, translating evidence into policy requires coordination among federal and provincial governments, immunization programme managers, clinicians, researchers, and community stakeholders (18–20). While surveillance studies quantify disease burden, they provide limited insight into how communities and decision-makers perceive a new vaccine or how evidence may be translated into future vaccine introduction.

Several Shigella vaccine candidates are currently in advanced stages of clinical development, increasing the likelihood of their future introduction into national immunization programmes (15, 21).

Evidence from LMICs suggests that decisions regarding Shigella vaccine prioritization depend not only on disease burden but also on programmatic feasibility, stakeholder engagement, and community confidence (21). Pakistan’s experience introducing the typhoid conjugate vaccine (TCV) similarly demonstrates the importance of institutional coordination, delivery planning, and community trust in achieving successful vaccine implementation (22–24). Consistent with this, WHO guidance emphasizes that successful vaccine introduction requires an understanding of community perceptions, stakeholder priorities, and health-system readiness alongside technical and epidemiological evidence (16, 25).

Little is known about how communities and key stakeholders in Pakistan perceive Shigella, interpret emerging surveillance evidence, and anticipate the introduction of a future Shigella vaccine.

Addressing this knowledge gap before vaccine licensure may help identify opportunities and barriers to implementation and inform early planning. We therefore explored perceptions of diarrhoeal disease and Shigella, factors influencing acceptance of a future Shigella vaccine, and stakeholder perspectives on vaccine prioritization and health-system readiness in Pakistan.

## Methods

### Study design and setting

We conducted a qualitative descriptive study between August 2024 and March 2025 to explore perceptions of diarrhoeal disease and Shigella, factors influencing acceptance of a future Shigella vaccine, and health-system readiness for its introduction into Pakistan’s routine immunization programme.

Focus group discussions (FGDs) were conducted with parents and caregivers of children under five years attending healthcare facilities participating in the Enterics for Global Health (EFGH) Shigella surveillance study in Karachi (13). These sites represent high-burden urban settings with ongoing diarrhoeal disease surveillance. Caregivers were recruited from healthcare facilities participating in the EFGH surveillance study and had recent experience with childhood diarrhoeal illness and healthcare seeking, enabling exploration of perceptions grounded in lived experiences of disease and care-seeking. FGDs explored perceptions of diarrhoeal diseases and vaccination and identified potential barriers and facilitators to the uptake of a future Shigella vaccine.

In-depth interviews (IDIs) were conducted with policymakers and program implementers from the Expanded Program on Immunization (EPI) across three provinces, Sindh, Khyber Pakhtunkhwa (KPK), Punjab, and the Federal Directorate of Immunization (FDI) Pakistan, program managers, pediatricians, school administrators, religious leaders, and community mobilizers. These stakeholders were selected to capture perspectives across different levels of the health system and community influence, including policy, program implementation, service delivery, and community engagement.

### Study population and sampling

A purposive sampling strategy was used to recruit participants across key stakeholder groups. Participants were selected to capture complementary perspectives on vaccine introduction from both demand and supply sides of the health system. Individuals were selected for their roles in immunization policy, program implementation, or community engagement, identified through institutional networks, and approached in person or by phone 16 IDIs were conducted with the stakeholders.

Caregivers of children under five years attending EFGH study sites were recruited to capture a range of perspectives on vaccination. Each FGD included 6-8 participants and 6 FGDs were conducted with the caregivers.

### Data collection

Data were collected using semi-structured interview guides developed in line with the study objectives, informed by the Health Belief Model, and adapted to the local context. FGDs and IDIs were conducted by trained qualitative researchers experienced in facilitating interviews and group discussions. All interviews and discussions were conducted in Urdu and audio-recorded with participants’ consent. Basic demographic characteristics of participants were collected using a short, structured form. Predefined sample sizes and study objectives guided data collection to ensure adequate depth and diversity of perspectives across participant groups.

FGDs were conducted at accessible community-based locations, including health centers and clinics, ensuring privacy and a quiet environment. Each session lasted approximately 60 minutes. Two facilitators were present: one moderated the discussion while the other took detailed field notes. Transportation was provided to participants as needed.

IDIs were conducted in private settings, including offices and EPI centers, using a flexible approach that allowed probing while maintaining consistency across interviews. Each interview lasted approximately 45-60 minutes.

Interviews and focus group discussions were conducted by NA, a male Research Manager with an MSc degree, and NM, a female Research Specialist with an MSc degree. Both researchers were based at Aga Khan University and had prior experience and training in qualitative data collection. Before data collection, participants were informed about the interviewers’ institutional affiliation, their role in the research team, the purpose of the study, and the reasons for conducting the research. The interviewers had no prior relationship with caregiver participants and were not involved in participants’ clinical care. Reflexive discussions were held throughout data collection and analysis to consider how the researchers’ professional backgrounds, prior experience, and assumptions might influence data collection and interpretation.

### Data management and analysis

All interviews were audio-recorded, transcribed verbatim in Urdu, and translated into English by trained bilingual research staff. Translations were reviewed for accuracy by a second team member by cross-checking transcripts against audio recordings. All data were anonymized and stored securely in password-protected files.

Data were analyzed thematically following Braun and Clarke’s six-step approach (26). A hybrid approach combining deductive and inductive analysis was used. The Health Belief Model informed the initial deductive coding framework, while inductive coding allowed additional concepts to emerge from the data.

A codebook was developed and applied independently by two researchers. Discrepancies were resolved through discussion and iterative refinement of codes and themes.

Rigor was enhanced using Lincoln and Guba’s criteria, including reflexivity to minimize bias, prolonged engagement with the data, and the use of verbatim quotations to support interpretation (27). Study procedures, participant selection, and analytic decisions were systematically documented. Reflexive field notes were maintained throughout data collection and analysis. Peer review of data collection tools and analytic outputs further enhanced credibility and dependability.

### Ethical considerations

Ethical approval was obtained from the Institutional Ethics Review Committee of Aga Khan University. Written informed consent was obtained from all participants prior to data collection. For participants unable to read or write, consent was obtained through thumbprint in the presence of a witness.

Participant confidentiality was preserved by assigning unique identification numbers. FGDs and interviews took place in private settings. No monetary incentives were provided; however, refreshments were offered following the interviews.

### Patient and public involvement

Patients and members of the public were not involved in the design of the study, development of the research questions, or interpretation of the findings. Caregivers participated as research participants through focus group discussions. The findings will be disseminated to relevant stakeholders through scientific publications and dissemination meetings.

## Results

A total of 16 IDIs and six FGDs were conducted. IDIs included policymakers, immunization programme managers, technical advisors, healthcare providers, education sector representatives, community mobilizers, and religious leaders. FGDs involved caregivers of children under five years attending EFGH surveillance sites. The findings are organized into four themes: (1) perceptions of diarrhoeal disease and Shigella, (2) factors shaping acceptance of a future Shigella vaccine, (3) evidence and health-system readiness for vaccine introduction, and (4) vaccine attributes, adoption, and sustainability. Participant characteristics are summarized in **Table la and lb**. Themes, subthemes, and illustrative quotations are presented in **Table 2**.

**Table 1a.** Characteristics of stakeholders participating in in-depth interviews (n = 16)

| Participant category | Number of participants | Description |
| --- | --- | --- |
| EPI programme managers | 4 | Provincial and Federal EPI representatives |
| Technical advisors on immunization | 4 | Immunization and vaccine programme experts from WHO and Gates Foundation |
| Provincial Ministry of Health | 2 | Representatives involved in disease surveillance and immunization campaigns |
| Healthcare providers (paediatricians) | 2 | Clinicians involved in child health care |
| School administrators | 2 | Education sector representatives involved in vaccine catch-up campaigns |
| Religious and Community leaders | 2 | Community engagement |
| **Total** | 16 | Multi-stakeholder perspectives on future Shigella vaccine introduction |

**Table 2.**
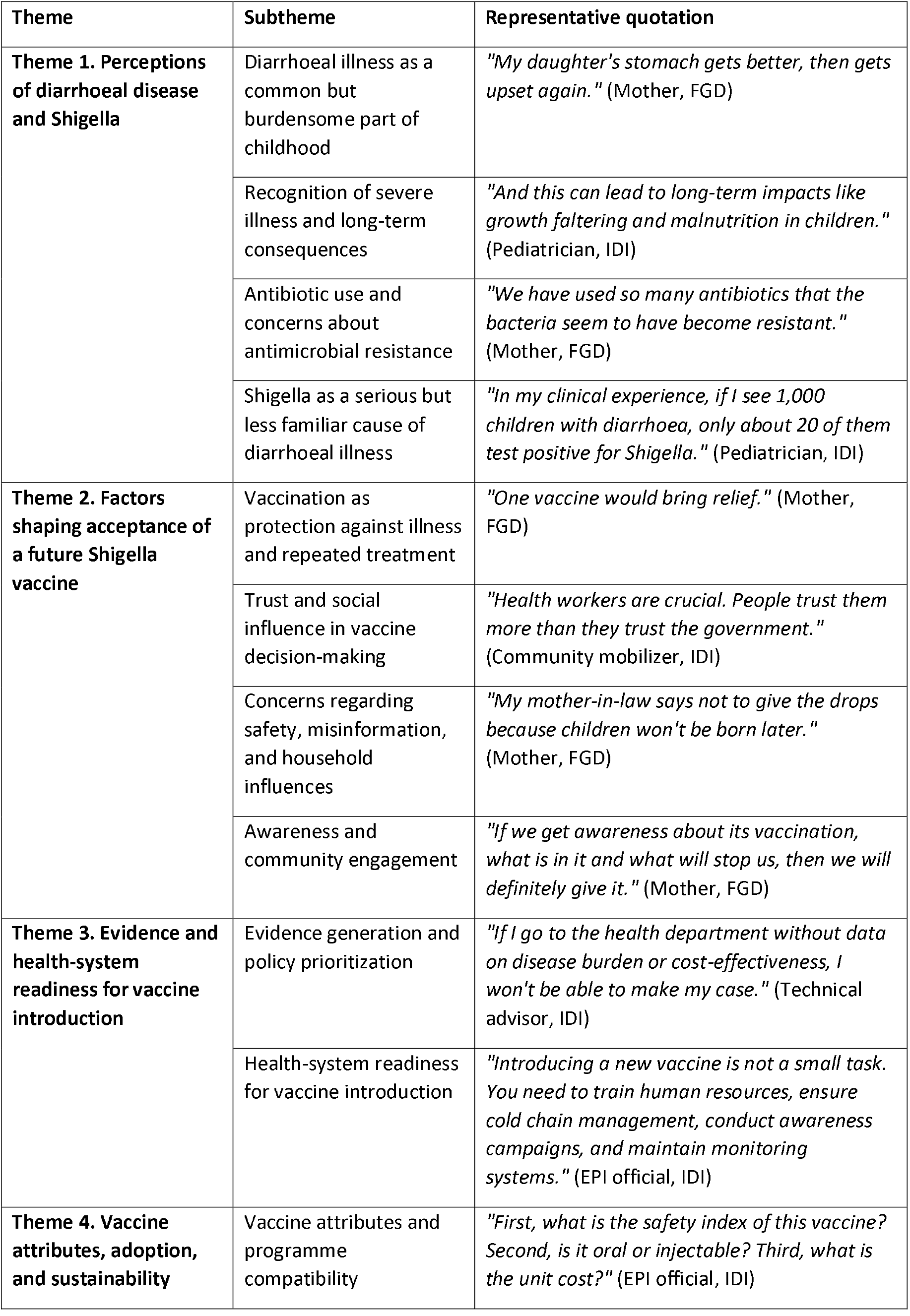
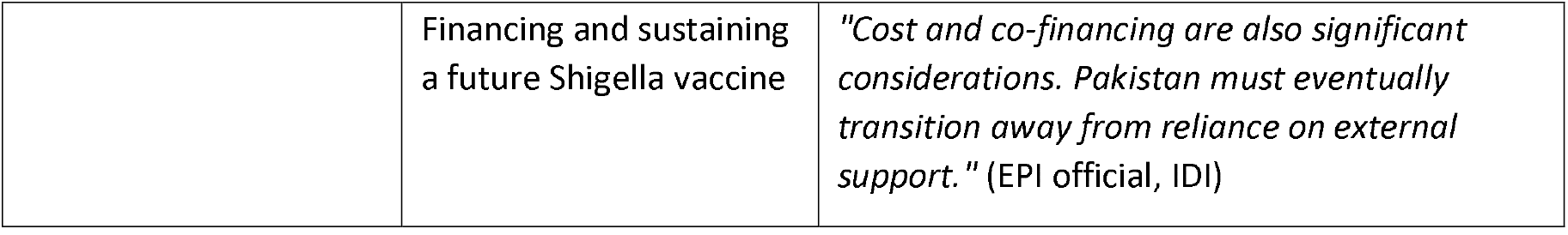
Themes, subthemes and representative quotations.

| Theme | Subtheme | Representative quotation |
| --- | --- | --- |
| **Theme 1. Perceptions of diarrhoeal disease and Shigella** | Diarrhoeal illness as a common but burdensome part of childhood | *"My daughter's stomach gets better, then gets upset again."* (Mother, FGD) |
|  | Recognition of severe illness and long-term consequences | *"And this can lead to long-term impacts like growth faltering and malnutrition in children."* (Pediatrician, IDI) |
|  | Antibiotic use and concerns about antimicrobial resistance | *"We have used so many antibiotics that the bacteria seem to have become resistant."* (Mother, FGD) |
|  | Shigella as a serious but less familiar cause of diarrhoeal illness | *"In my clinical experience, if I see 1,000 children with diarrhoea, only about 20 of them test positive for Shigella."* (Pediatrician, IDI) |
| **Theme 2. Factors shaping acceptance of a future Shigella vaccine** | Vaccination as protection against illness and repeated treatment | *"One vaccine would bring relief."* (Mother, FGD) |
|  | Trust and social influence in vaccine decision-making | *"Health workers are crucial. People trust them more than they trust the government."* (Community mobilizer, IDI) |
|  | Concerns regarding safety, misinformation, and household influences | *"My mother-in-law says not to give the drops because children won't be born later."* (Mother, FGD) |
|  | Awareness and community engagement | *"If we get awareness about its vaccination, what is in it and what will stop us, then we will definitely give it."* (Mother, FGD) |
| **Theme 3. Evidence and health-system readiness for vaccine introduction** | Evidence generation and policy prioritization | *"If I go to the health department without data on disease burden or cost-effectiveness, I won't be able to make my case."* (Technical advisor, IDI) |
|  | Health-system readiness for vaccine introduction | *"Introducing a new vaccine is not a small task. You need to train human resources, ensure cold chain management, conduct awareness campaigns, and maintain monitoring systems."* (EPI official, IDI) |
| **Theme 4. Vaccine attributes, adoption, and sustainability** | Vaccine attributes and programme compatibility | *"First, what is the safety index of this vaccine? Second, is it oral or injectable? Third, what is the unit cost?"* (EPI official, IDI) |
|  | Financing and sustaining a future Shigella vaccine | *"Cost and co-financing are also significant considerations. Pakistan must eventually transition away from reliance on external support."* (EPI official, IDI) |

### Theme 1: Perceptions of diarrhoeal disease and Shigella

Participants described childhood diarrhoeal illness as a common childhood illness that placed a considerable burden on children and their families. While diarrhoea itself was widely recognized, most caregivers were unfamiliar with Shigella before it was introduced during the discussions.

Caregivers and stakeholders also reflected on the severity of diarrhoeal illness, concerns about antimicrobial resistance, and the potential role of preventive measures.

### Diarrhoeal illness as a common but burdensome part of childhood

Caregivers frequently described diarrhoea as one of the most common illnesses affecting young children. Many spoke about repeated episodes during early childhood and the ongoing burden of caring for children who became ill repeatedly. Participants commonly linked these recurrent illnesses to poor water quality, inadequate sanitation and hygiene, and children’s everyday exposure to environmental risks.

> *“They suffer more from diarrhea. Children put everything in their mouths, and because of this, every few days we face one problem or another. Whether it’s heat-related diarrhea or something else, we are constantly worried*.*” (Mother, FGD)*

Similarly, another caregiver reflected on the persistent nature of diarrhoeal illness:

> *“My daughter’s stomach gets better, then gets upset again*.*” (Mother, FGD)*

Caregivers commonly attributed diarrhoeal illness to contaminated food, unsafe drinking water, poor sanitation, inadequate handwashing, and other environmental exposures, rarely referring to specific infectious pathogens. Many described diarrhoea as difficult to fully prevent because children are continually exposed to these risks in their everyday environment. Reflecting a similar perspective, one stakeholder emphasized that the persistence of diarrhoeal disease extends beyond healthcare and is closely linked to environmental conditions:

> *“Diarrhea has persisted for generations, new cohorts of children face the same risks as the previous ones. The issue is not solely medical,it involves sanitation, hygiene, and water supply*.*” (Health ministry official, IDI)*

### Recognition of severe illness and long-term consequences

Participants described diarrhoeal illness as becoming particularly concerning when children developed dehydration, bloody diarrhoea, or required hospitalization. Caregivers frequently sought medical care when symptoms worsened or failed to improve.

> *“When the condition got really bad with fever and excessive diarrhea, that’s when I brought them here, and the doctors put them on an IV* ^*”*^ *(Mother, FGD)*

Beyond the acute illness, caregivers described children becoming weak, losing weight, or eating poorly after repeated episodes of diarrhoea. Healthcare providers further linked recurrent enteric infections to malnutrition, micronutrient deficiencies, growth faltering, and stunting.

> *“And this can lead to long-term impacts like growth faltering and malnutrition in children*.*” (Pediatrician, IDI)*

### Antibiotic use and concerns about antimicrobial resistance

Concerns about antimicrobial resistance emerged across both caregiver and stakeholder groups. Caregivers frequently described treating diarrhoeal illness with antibiotics, often using medicines that had been prescribed previously or were already available at home. Flagyl (metronidazole) and Entamizole were repeatedly mentioned as medicines used at the onset of illness, and several participants described keeping these medicines at home before seeking formal care.

> *“So Ifirst gave them Flagyl to try to control it. I took the Flagyl along with Entamizole; it stopped for two days, but then it started again*. ^*”*^ *(Mother, FGD)*

Some participants felt that these treatments had become less effective over time.

> *“We have used so many antibiotics that the bacteria seem to have become resistant*.*” (Mother, FGD)*

Healthcare providers described similar concerns and explained that limited access to diagnostic testing often resulted in empirical antibiotic treatment.

> *“Often, we don’t get proper cooperation from patients for stool cultures. We usually go by symptoms and start them on antibiotics*.*” (Pediatrician, IDI)*

One pediatrician also reflected on how commonly metronidazole was used in household management of diarrhoeal illness:

> *“In many households, it’s treated like a national ‘diarrhea drink*.*’ People assume it is the solution to every case of diarrhea*. ^*”*^ *(Pediatrician, IDI)*

### Shigella as a serious but less familiar cause of diarrhoeal illness

Most caregivers were unfamiliar with Shigella before it was introduced during the discussions. After learning that it could cause bloody diarrhoea, participants generally regarded it as a serious illness requiring prompt treatment. Healthcare providers similarly distinguished Shigella from more routine episodes of diarrhoea and emphasized its potential to cause more severe disease.

One pediatrician contrasted the findings of *Shigella* surveillance conducted in Pakistan with his own clinical experience:

> *“Data suggests that 35% of 14,000 samples were positive for Shigella [participant referring to Shigella surveillance study]. As a pediatrician, I find that number quite high. In my clinical experience, if I see 1,000 children with diarrhoea, only about 20 of them test positive for Shigella. So, the actual incidence is relatively low*.*” (Pediatrician, IDI)*

Some clinicians also emphasized that vaccination alone would not be sufficient to reduce the burden of Shigella and highlighted the continued importance of improving water, sanitation, and hygiene.

> *“Since Shigella is transmitted through the fecal-oral route, any vaccination effort must also be accompanied by improvements in water, sanitation, and hygiene (WASH). A vaccine alone won’t be enough-we need an integrated approach*.*” (EPI official, IDI)*

### Theme 2. Factors shaping acceptance of a future Shigella vaccine

Although awareness of Shigella was generally low, caregivers and stakeholders were broadly supportive of vaccination to protect children from illness. However, acceptance of a future Shigella vaccine was rarely described as unconditional. Instead, participants highlighted several factors that would influence their decisions, including trust, perceptions of vaccine safety, and access to clear, reliable information.

### Vaccination as protection against illness and repeated treatment

Caregivers frequently described vaccination as a practical way to protect children from illness and reduce the need for repeated treatment. Many viewed vaccination as preferable to giving medicines repeatedly, particularly when children experienced recurrent episodes of diarrhoeal illness.

> *“It would benefit the children; it’s a good thing. We wouldn’t have to give them medicine over and over again. One vaccine would bring relief” (Mother, FGD)*

Stakeholders also expressed positive views about vaccination and described it as an important way to prevent illness, provided that the vaccine was shown to be safe and effective.

### Trust and social influence in vaccine decision-making

Stakeholders described trusted individuals and existing community networks as important channels for communicating information about vaccination. Healthcare providers, community health workers, religious leaders, community mobilizers, and other respected local figures were frequently identified as trusted sources of information for families considering vaccination.

> *“Health workers are crucial. People trust them more than they trust the government. Mothers, in particular, tend to listen because they’re directly involved in their children’s nutrition and health*.*” (Community mobilizer, IDI)*

Reflecting on the COVID-19 vaccination programme, one EPI official described how information was shared through trusted local health workers and spread by word of mouth within the community.

> *“We have strong, trust-based relationships with them, and they are well-connected in the community …That word-of-mouth approach helped us spread the message effectively*.*” (EPI official, IDI)*

### Concerns regarding safety, misinformation, and household influences

Despite generally positive attitudes toward vaccination, participants described concerns about vaccine safety, children becoming ill after vaccination, and misinformation circulating within their communities. Perceived side effects, including fever, pain, and other illnesses following vaccination, were commonly discussed and sometimes contributed to hesitancy. Rumors about infertility and vaccine safety were also reported across several communities.

> *“My mother-in-law says not to give the drops because children won’t be born later*.*” (Mother, FGD)*

Stakeholders similarly described how misinformation spread rapidly through social media and informal conversations, making it difficult to address once widely accepted.

> *“Once something becomes popular, social media is enough to spread it. These ideas enter people’s minds, and it becomes very difficult to remove them*.*” (Religious leader, IDI)*

Caregivers also described how vaccination decisions were influenced by experiences and advice shared within families and social circles. Husbands, mothers-in-law, relatives, and friends were frequently mentioned as shaping attitudes toward vaccination, particularly following reports of perceived side effects or illness after vaccination.

### Awareness and community engagement

Caregivers and stakeholders described the need for clear information and community engagement before a future Shigella vaccine could be widely accepted. Some caregivers said they would be willing to vaccinate their children but wanted clear information about the disease, the vaccine, its benefits, and possible side effects before making a decision.

> *“If we get awareness about its vaccination, what is in it and what will stop us, then we will definitely give* it.” *(Mother, FGD)*

Stakeholders described several approaches for engaging communities, including healthcare providers, lady health workers, schools, religious leaders, and community mobilizers. They also noted that raising awareness would be particularly important because most communities were unfamiliar with Shigella.

> *“The best thing is for the person in front of you to be ready to listen. Whether they believe or not is one thing, but within a week or so, you can turn their refusal into acceptance*.*” (Religious leader, IDI)*

### Theme 3. Evidence and health-system readiness for vaccine introduction

Stakeholders discussed a range of considerations related to the introduction of a future Shigella vaccine in Pakistan. Their discussions focused on the evidence needed to inform policy decisions and the capacity of the existing immunization system to support vaccine introduction and delivery.

### Evidence generation and policy prioritization

Stakeholders emphasized that decisions regarding the introduction of new vaccines should be guided by evidence on disease burden. Participants highlighted the importance of local- and national-level data, including surveillance evidence, antimicrobial resistance patterns, cost-effectiveness, and the potential contribution of Shigella to malnutrition and growth faltering, to inform policy decisions.

> *“If I go to the health department without data on disease burden or cost-effectiveness, I won’t be able to make my case*.*” (Technical advisor, IDI)*

Several stakeholders also highlighted the importance of generating evidence from across Pakistan. While acknowledging the value of existing surveillance activities, participants noted that findings from a limited number of sites would be insufficient to inform national policy decisions.

> *“We need more research and concrete evidence to convince policymakers. If the data is limited to one city or one district, it’s harder to implement policies on a national level*.*” (FDI official, IDI)*

Technical advisor and EPI officials described how decisions about new vaccines are made alongside other public health priorities. Polio, measles, dengue, malaria, and typhoid were frequently mentioned as conditions that currently receive greater attention and resources. While some participants considered the available evidence on Shigella sufficient to begin planning discussions, others felt that broader surveillance, burden estimates, and cost-effectiveness data would still be needed before a future Shigella vaccine could be prioritized.

> *“There are many competing priorities, and policymakers naturally focus on diseases that have stronger visibility and stronger data*.*” (Technical advisor, IDI)*

### Health-system readiness for vaccine introduction

Stakeholders generally viewed Pakistan’s existing EPI as a useful foundation for introducing future vaccines. Existing delivery systems, cold-chain infrastructure, vaccination centres, community outreach mechanisms, and experience with previous vaccine introductions were described as important strengths.

> *“We already have the infrastructure; the challenge is how effectively we use it*.*” (EPI official, IDI)*

Several participants referred to Pakistan’s experience with rotavirus and typhoid conjugate vaccines as examples of how the immunization system had previously adapted to new vaccines. At the same time, stakeholders emphasized that introducing a future Shigella vaccine would still require careful preparation, including training healthcare workers, maintaining cold-chain systems, conducting awareness campaigns, strengthening monitoring mechanisms, and preparing communities.

> *“Introducing a new vaccine is not a small task. You need to train human resources, ensure cold chain management, conduct awareness campaigns, and maintain monitoring systems” (EPI official, IDI)*.
>
> Stakeholders described coordination between researchers, policymakers, immunization managers, healthcare providers, community mobilizers, educators, and religious leaders. These actors were described as important for translating evidence into implementation and for supporting awareness and acceptance at the community level.

### Theme 4. Vaccine attributes, adoption, and sustainability

Stakeholders discussed several characteristics they considered important for a future Shigella vaccine, as well as the financial and programmatic considerations that could influence its adoption and long term sustainability in Pakistan.

### Vaccine attributes and programme compatibility

Stakeholders discussed several characteristics they considered important when reflecting on a future Shigella vaccine. Safety and effectiveness were frequently identified as key considerations, alongside the duration of protection, coverage against locally circulating strains, route of administration, and the incorporation of the vaccine into Pakistan’s existing immunization schedule.

> *“First, what is the safety index of this vaccine? Second, is it oral or injectable? Third, what is the unit cost?” (EPI official, IDI)*

Participants also discussed the importance of information on locally circulating strains, vaccine performance, dosing schedules, and duration of protection before decisions about vaccine introduction could be made.

> *“We need to determine whether strain mapping has been done in Pakistan*.*” (EPI official, IDI)*

Stakeholders also discussed how the route of administration could influence programme delivery and community acceptance. Some participants felt that an oral vaccine might be easier to administer and more acceptable to caregivers, while others noted that the vaccine should fit within existing EPI contact points and avoid unnecessary additional visits where possible.

> *“If the Shigella vaccine is oral, there will be no issue. Injectable vaccines, however, do face resistance*.*” (EPI official, IDI)*

Several EPI officials and technical advisors emphasized that a future Shigella vaccine should fit within existing EPI contact points rather than require additional visits. Some also discussed combination vaccines as a way to reduce the number of injections, facilitate integration into the existing immunization schedule, and make delivery easier for caregivers and vaccinators.

> *“You cannot just add it randomly. Each child already comes in six times for EPI vaccines. If we start requiring seven or eight visits, it becomes impractical*.*” (EPI official, IDI)*

### Financing and sustaining a future Shigella vaccine

Stakeholders described financing as an important consideration for the introduction of a future Shigella vaccine. Participants frequently referred to the roles of Gavi, WHO, UNICEF, and other development partners in supporting vaccine procurement and implementation, while noting that long-term sustainability would require increasing government ownership and domestic financing.

> *“Cost and co-financing are also significant considerations. Pakistan must eventually transition away from reliance on external support*.*” (EPI official, IDI)*

Several stakeholders discussed the financial challenges of introducing and maintaining new vaccines. Affordability, available resources, and long-term government commitment were commonly described as important considerations when discussing whether a future Shigella vaccine could be sustained within the national immunization programme.

> *“The introduction of a new vaccine depends on several factors: the government’s acceptance, the disease burden, and, most importantly, financial support. With support, the government’s sustainability also plays a key role*.*” (FDI official, IDI)*

. One stakeholder referred to a previous proposal for local vaccine manufacturing, noting that it was not pursued because purchasing vaccines was considered less costly than investing in domestic production capacity.

## Discussion

This study explored community and stakeholder perspectives on the potential introduction of a future Shigella vaccine in Pakistan. Although awareness of Shigella was limited, participants generally viewed vaccination positively. More importantly, stakeholders consistently described vaccine introduction as depending not only on community acceptance but also on the availability of locally relevant evidence, competing policy priorities, and sustainable implementation. These findings suggest that the principal challenge for Shigella vaccine introduction in Pakistan lies not simply in generating demand, but in strengthening the evidence base and creating the conditions necessary for policy adoption.

Our study findings suggest that stakeholders distinguished between recognizing Shigella as an important cause of childhood diarrhoeal disease and having sufficient evidence to justify its prioritization for vaccine introduction. Although clinicians and technical stakeholders acknowledged its contribution to antimicrobial resistance, malnutrition, and growth faltering, they consistently emphasized the need for stronger locally generated evidence on disease burden, surveillance, and cost-effectiveness before a future vaccine could be considered. This finding is consistent with Fleming et al., who reported that stakeholders in several low- and middle-income countries identified disease burden, antimicrobial resistance, and economic evidence as central considerations for the future introduction of a Shigella vaccine (21). Similarly, qualitative studies and subsequent reviews of national vaccine decision-making have consistently shown that disease burden, economic evidence, competing health priorities, and health-system capacity are central to decisions about introducing new vaccines in LMICs (28–30). In Pakistan, the multi-country Shigella surveillance study has generated important epidemiological, microbiological, and economic evidence to support Shigella vaccine development and inform future vaccine policy (13, 15, 31). However, stakeholders in our study considered these data an important foundation but emphasized the need for broader, nationally representative evidence to inform vaccine introduction decisions. Together, these findings highlight the importance of expanding high-quality surveillance and generating nationally relevant epidemiological and economic evidence to support the future introduction of a Shigella vaccine in Pakistan.

Although participants generally expressed positive attitudes toward vaccination, acceptance of a future Shigella vaccine was rarely described as uncnditional. Instead, caregivers emphasized the importance of understanding the vaccine, discussing their concerns, and receiving recommendations from trusted individuals before deciding to vaccinate their children. These findings are consistent with the WHO Behavioural and Social Drivers (BeSD) framework, which identifies confidence, social influences, and trusted provider recommendations as important determinants of vaccine uptake (32). Similar observations have been reported during the introduction of the typhoid conjugate vaccine in Pakistan, where healthcare providers and community stakeholders played an important role in addressing concerns and supporting vaccine acceptance (23, 24). This finding challenges the simplistic binary acceptance-versus-hesitancy frameworks, instead revealing a more nuanced decision-making process requiring information, trust, and social validation.

Stakeholders generally viewed Pakistan’s existing EPI as a strong foundation for introducing a future Shigella vaccine. Their confidence was shaped by Pakistan’s experience introducing rotavirus vaccine into the routine EPI schedule and typhoid conjugate vaccine through outbreak response, catch-up campaigns, and routine immunization integration. Pakistan’s TCV experience is particularly relevant, as local evidence on XOR typhoid supported vaccine introduction, followed by a catch-up campaign in Sindh and routine EPI integration with Gavi support (22–24, 33, 34). Rotavirus vaccine introduction similarly demonstrates Pakistan’s ability to incorporate a new diarrhoeal disease vaccine into the national immunization programme (35). These experiences reinforce WHO guidance that successful vaccine introduction depends not only on disease burden but also on operational readiness, financing, supply systems, implementation capacity, and long-term sustainability (16). Similar priorities have also been highlighted during international discussions on the development of future Shigella vaccines, particularly regarding schedule compatibility and product characteristics (36).

These findings have important implications for preparing for the future introduction of a Shigella vaccine in Pakistan. Expanding surveillance beyond sentinel sites to generate geographically representative data on disease burden, antimicrobial resistance, and costs will be critical for informing national policy decisions. Importantly, our findings suggest that communication strategies for Shigella vaccine introduction must transcend traditional information dissemination approaches to create meaningful dialogue opportunities through established community trust networks. This is particularly critical given the limited baseline awareness of Shigella observed across all participant categories, necessitating comprehensive education campaigns that build understanding while addressing potential concerns proactively. These priorities are consistent with WHO guidance on new vaccine introduction and Gavi 6.0, which emphasize country ownership, sustainability, equity, and stronger immunization systems (16, 37).

This study has several strengths. By incorporating perspectives from caregivers, healthcare providers, policymakers, programme managers, technical experts, and community stakeholders, we captured views across multiple levels of the health system. Including stakeholders from provincial EPI and the FDI provided insights into policy and programmatic considerations beyond community acceptance. However, there are several limitations. Caregivers were recruited from EFGH surveillance sites in urban and peri-urban Karachi and may not reflect the experiences of populations in rural areas or other parts of Pakistan. As awareness of Shigella was generally low, participants received a brief explanation of the pathogen during discussions, which may have influenced their views on a future vaccine. Finally, as with all qualitative studies, these findings provide in-depth insights into stakeholder perspectives but are not intended to be statistically generalizable.

## Conclusion

Although childhood diarrhoeal disease was widely recognized, awareness of Shigella as a specific pathogen remained limited. Participants were generally receptive to a future Shigella vaccine, but acceptance depended on trust, confidence in vaccine safety, and access to reliable information. Stakeholders viewed vaccine introduction as feasible within Pakistan’s existing immunization system, provided it is supported by stronger locally generated evidence, sustained policy commitment, and early implementation planning. Together, these findings suggest that strengthening the evidence base while engaging trusted community networks and building on Pakistan’s established EPI platform will be critical to preparing for the successful introduction of a future Shigella vaccine.

## Data Availability

All data produced in the present study are available upon reasonable request to the authors

## Author Contributions

Naveed Ahmed (NA) conceptualized the study, developed the interview guides, funding acquisition, coordinated data collection, conducted the formal analysis and interpretation of the data, and drafted the original manuscript. Contributed to securing funding.

Nasreen Mithani **(NM)**, Zubair Latif (ZL), contributed to data collection, data management, transcriptions, and interpretation of the findings. They also reviewed the manuscript.

Irum Fatima (IF), and Irfan Gaffar (IG) contributed to reviewing the manuscript.

Mohammad Tahir Yousafzai (TY) contributed to study conceptualization, methodology, interpretation of findings, and critically reviewed the manuscript for important intellectual content.

Farah Naz Qamar (FN) conceived and supervised the study, secured funding, provided overall scientific oversight, contributed to study design and interpretation of the findings, and critically reviewed the manuscript for important intellectual content.

All authors reviewed and approved the final version of the manuscript.

## Acknowledgments

We sincerely thank all study participants for generously sharing their time, experiences, and perspectives, which made this research possible.

We acknowledge the financial support provided through the Enterics for Global Health (EFGH) seed grant funded by the University of Washington and the Gates Foundation.

We are grateful to the EFGH coordination team at the University of Washington, particularly Sonia Rao, Kirk Tickell, and Patricia B. Pavlinac, for their continuous guidance, encouragement, and support throughout this project.

We extend our special thanks to Arianna R. Means from the University of Washington for her thoughtful and constructive feedback on the development of the qualitative interview guides.

## Data Sharing

Deidentified data will be made available after the publication of this manuscript upon reasonable request to the author.

---

Sociodemographic and household characteristics of caregivers participating in focus group discussions (n=46)

| Characteristic* | n (%) |
| --- | --- |
| **Caregiver age, years** |  |
| Mean ± SD | 26.5 ± 4.7 |
| Median (IQR) | 26.5 (24.0–29.0) |
| **Caregiver employment status** |  |
| Not employed | 2 (4.3) |
| Housewife | 44 (95.7) |
| **Number of children aged <5 years in the household** |  |
| 1 | 18 (39.1) |
| 2 | 20 (43.5) |
| $\geq 3$ | 8 (17.4) |
| **Main source of drinking water** |  |
| Piped water into dwelling | 36 (78.3) |
| Piped into compound, yard or plot | 2 (4.3) |
| Filtered water kiosk | 7 (15.2) |
| Bottled water | 1 (2.2) |
| **Type of toilet facility** |  |
| Improved: flush or pour-flush to piped sewer | 41 (89.1) |
| Unimproved: bucket toilet | 5 (10.9) |
| **Household wealth quintile** |  |
| Poorest | 11 (23.9) |
| Poor | 16 (34.8) |
| Middle | 4 (8.7) |
| Rich | 11 (23.9) |
| Richest | 4 (8.7) |

Data presented as Mean ± SD and n (%)
\*Each FGD consisted of 6–8 participants

## References

1. WHO. Diarrhoeal disease 2024 [cited 2026 09 Feb]. Available from: https://www.who.int/news-room/fact-sheets/detail/diarrhoealdisease#:~:text=Key%20facts,of%20all%20diarrhoea%2Dassociated%20deaths.

2. Troeger C, Blacker BF, Khalil IA, Rao PC, Cao S, Zimsen SR, et al. Estimates of the global, regional, and national morbidity, mortality, and aetiologies of diarrhoea in 195 countries: a systematic analysis for the Global Burden of Disease Study 2016. The Lancet Infectious Diseases. 2018;18(11):1211–28.

3. Zhu H-Y, Xu F, Zhao W-Z, Wang H-X, Wang H-G. The global burden of childhood diarrhea and its epidemiological characteristics from 1990 to 2021. Frontiers in Pediatrics. 2025;13:1656234.

4. Rahmat ZS, Zubair A, Abdi I, Humayun N, Arshad F, Essar MY. The rise of diarrheal illnesses in the children of Pakistan amidst COVID-19: A narrative review. Health Science Reports. 2023;6(1):e1043.

5. Yousafzai MT, Cornick J, Yori PP, Hossain MJ, Keita AM, Atlas HE, et al. Incidence and Antimicrobial Resistance Of Shigella-Attributable Diarrhea in Young Children: Results from the Multi-Country Enterics for Global Health (Efgh) Shigella Surveillance Study. 2025.

6. Kotloff KL, Nataro JP, Blackwelder WC, Nasrin D, Farag TH, Panchalingam S, et al. Burden and aetiology of diarrhoeal disease in infants and young children in developing countries (the Global Enteric Multicenter Study, GEMS): a prospective, case-control study. The lancet. 2013;382(9888):209–22.

7. Platts-Mills JA, Liu J, Rogawski ET, Kabir F, Lertsethtakarn P, Siguas M, et al. Use of quantitative molecular diagnostic methods to assess the aetiology, burden, and clinical characteristics of diarrhoea in children in low-resource settings: a reanalysis of the MAL-ED cohort study. The Lancet Global Health. 2018;6(12):e1309–e18.

8. Khalil IA, Troeger C, Blacker BF, Rao PC, Brown A, Atherly DE, et al. Morbidity and mortality due to shigella and enterotoxigenic Escherichia coli diarrhoea: the Global Burden of Disease Study 1990–2016. The Lancet infectious diseases. 2018;18(11):1229–40.

9. Libby TE, Delawalla MLM, Al-Shimari F, MacLennan CA, Vannice KS, Pavlinac PB. Consequences of Shigella infection in young children: a systematic review. Int J Infect Dis. 2023;129:78–95.

10. Puett C, Anderson JD, Bagamian KH, Muhib F, Scheele S, Hausdorff WP, Pecenka C. Projecting the long-term economic benefits of reducing Shigella-attributable linear growth faltering with a potential vaccine: a modelling study. The Lancet Global Health. 2023;11(6):e892–e902.

11. CDC. Antimicrobial Resistance and Shigella Infections 2024 [cited 2026 Feb 09,]. Available from: https://www.cdc.gov/shigella/treatment/antimicrobial-resistance-and-shigellainfections.html#:~:text=Key%20points,who%20have%20sex%20with%20men.

12. Studies NIoP. Pakistan demographic and health survey 2017–18. NIPS/Pakistan and ICF Islamabad, Pakistan; 2019.

13. Atlas HE, Conteh B, Islam MT, Jere KC, Omore R, Sanogo D, et al., editors. Diarrhea case surveillance in the Enterics for Global Health Shigella surveillance study: epidemiologic methods. Open forum infectious diseases; 2024: Oxford University Press US.

14. Kotloff KL, Nasrin D, Blackwelder WC, Wu Y, Farag T, Panchalingham S, et al. The incidence, aetiology, and adverse clinical consequences of less severe diarrhoeal episodes among infants and children residing in low-income and middle-income countries: a 12-month case-control study as a follow-on to the Global Enteric Multicenter Study (GEMS). The Lancet Global Health. 2019;7(5):e568–e84.

15. Vannice K, MacLennan CA, Long J, Steele AD, editors. Optimizing vaccine trials for enteric diseases: the Enterics for Global Health (EFGH) Shigella surveillance study. Open Forum Infectious Diseases; 2024: Oxford University Press US.

16. Organization WH. Principles and considerations for adding a vaccine to a national immunization programme: from decision to implementation and monitoring. 2014.

17. Donohue JF, Elborn JS, Lansberg P, Javed A, Tesfaye S, Rugo H, et al. Bridging the “know-do” gaps in five non-communicable diseases using a common framework driven by implementation science. Journal of healthcare leadership. 2023:103–19.

18. Nishtar S, Boerma T, Amjad S, Alam AY, Khalid F, ul Haq I, Mirza YA. Pakistan’s health system: performance and prospects after the 18th Constitutional Amendment. The Lancet. 2013;381(9884):2193–206.

19. Zaidi S, Bigdeli M, Aleem N, Rashidian A. Access to essential medicines in Pakistan: policy and health systems research concerns. PloS one. 2013;8(5):e63515.

20. Haq Z, Shaikh BT, Tran N, Hafeez A, Ghaffar A. System within systems: challenges and opportunities for the Expanded Programme on Immunisation in Pakistan. Health research policy and systems. 2019;17(1):51.

21. Fleming JA, Gurley N, Knudson S, Kabore L, Bawa JT, Dapaah P, et al. Exploring Shigella vaccine priorities and preferences: Results from a mixed-methods study in low-and middle-income settings. Vaccine: X. 2023;15:100368.

22. Batool R, Qureshi S, Haq Z, Yousafzai MT, Salam RA, Ali R, et al. Coverage survey of typhoid conjugate vaccine among children aged 6 months to 15 years in an urban slum settlement of Lyari Town Karachi, Pakistan. Plos one. 2023;18(8):e0289582.

23. Batool R, Yousafzai MT, Qureshi S, Muhammad S, Qazi I, Sadaf T, et al. Parental acceptance of typhoid conjugate vaccine for children aged 6 months to 15 years in an outbreak setting of Lyari Town Karachi, Pakistan. Vaccine. 2023;41(37):5376–82.

24. Qamar FN, Batool R, Qureshi S, Ali M, Sadaf T, Mehmood J, et al. Strategies to improve coverage of typhoid conjugate vaccine (TCV) immunization campaign in Karachi, Pakistan. Vaccines. 2020;8(4):697.

25. Federal Directorate of Immunization P. National Immunization Policy 2022. 2022.

26. Braun V, Clarke V. Using thematic analysis in psychology. Qualitative research in psychology. 2006;3(2):77–101.

27. Lincoln YS, Guba EG. Criteria for Assessing Naturalistic Inquiries as Reports. 1988.

28. Guillaume D, Meyer D, Waheed D-e-N, Schlieff M, Muralidharan K, Chou VB, Limaye R. Factors influencing the prioritization of vaccines by policymakers in low-and middle-income countries: a scoping review. Health policy and planning. 2023;38(3):363–76.

29. Donadel M, Panero MS, Ametewee L, Shefer AM. National decision-making for the introduction of new vaccines: A systematic review, 2010–2020. Vaccine. 2021;39(14):1897–909.

30. Burchett HE, Mounier-Jack S, Griffiths U, Biellik R, Ongolo-Zogo P, Chavez E, et al. New vaccine adoption: qualitative study of national decision-making processes in seven low- and middle-income countries. Health policy and planning. 2012;27(suppl_2):ii5–ii16.

31. Ahmed N, Yousafzai MT, Naz Qamar F, editors. The Enterics for Global Health (EFGH) Shigella surveillance study in Pakistan. Open Forum Infectious Diseases; 2024: Oxford University Press US.

32. mondiale de la Santé O, Organization WH. Understanding the behavioural and social drivers of vaccine uptake WHO position paper–May 2022–Comprendre les facteurs comportementaux et sociaux de l’adoption des vaccins Note de synthèse de l’OMS–mai 2022. Weekly Epidemiological Record= Relevé épidémiologique hebdomadaire. 2022;97(20):209–24.

33. Yousafzai MT, Karim S, Qureshi S, Kazi M, Memon H, Junejo A, et al. Effectiveness of typhoid conjugate vaccine against culture-confirmed Salmonella enterica serotype Typhi in an extensively drug-resistant outbreak setting of Hyderabad, Pakistan: a cohort study. The Lancet Global Health. 2021;9(8):e1154–e62.

34. Batool R, Yousafzai MT, Qureshi S, Ali M, Sadaf T, Mehmood J, et al. Effectiveness of typhoid conjugate vaccine against culture-confirmed typhoid in a peri-urban setting in Karachi: a case-control study. Vaccine. 2021;39(40):5858–65.

35. Ali SA, Sultana S, Riaz A, Yousafzai MT, Hotwani A, Kabir F, et al. Effectiveness of monovalent rotavirus vaccine among young children in Pakistan: a test-negative case-control evaluation. Vaccine. 2026;81:128555.

36. Riddle MS, Bourgeois AL, Clifford A, Jeon S, Giersing BK, Jit M, et al. Challenges and opportunities in developing a Shigella-containing combination vaccine for children in low-and middle-income countries: Report of an expert convening. Vaccine. 2023;41(16):2634–44.

37. GAVI. Phase VI (2026–2030) 2024 [Available from: https://www.gavi.org/our-work/strategy/phase-6-2026-2030.

